# Major Depressive Disorder as a risk factor of neuropsychiatric symptoms in normal and pathological aging, and associations with cognitive performances

**DOI:** 10.1101/2022.05.04.22274088

**Authors:** Ronat Lucas, NACC Group, Hanganu Alexandru

**Affiliations:** Centre de Recherche de l’Institut Universitaire de Gériatrie de Montréal, Montréal, Québec, Canada; Faculté de Médecine, Département de Médecine, Université de Montréal, Québec, Canada; Faculté des Arts et des Sciences, Département de Psychologie, Université de Montréal, Québec, Canada

**Keywords:** Major Depressive Disorder, Cognitive Decline, Neuropsychiatric symptoms, Alzheimer’s Disease

## Abstract

**Objectives:** The diagnosis of Major Depressive Disorder (MDD) is based on the DSM-V criteria and is established by a clinician. It allows quantifying depression based on clinical criteria. As such, MDD differs from other types of depressions quantified based on subjective scales. Here, we evaluated the MDD risk factor on other neuropsychiatric symptoms (NPS) as well as MDD association with cognitive performance in Alzheimer’s disease (AD), Mild Cognitive Impairment (MCI) and Healthy Controls (CH).

**Participants:** Data of 208 patients with AD, 291 patients with MCI and 647 HC was extracted from the *National Alzheimer’s Coordinating Center* database. Each included participant was assessed by a physician for the MDD criteria, underwent an evaluation of NPS using the NeuroPsychiatric Inventory, and a comprehensive cognitive assessment. Participants were classified in those with- and without MDD. We performed logistic regression and a MANCOVA models respectively with NPS and cognitive performance as variables of interest and MDD as fixed factors within each group. The MANCOVA was controlled for the effects of age, sex, and education.

**Results:** MDD increased the risk for psychotic, affective and behavioral NPS in MCI, and affective/behavioral NPS in CH and AD. Also, MCI with MDD had lower performance on selective attention and mental flexibility.

**Conclusions:** MDD seems to increase the probability for a higher prevalence of NPS in all groups (CN, MCI and AD). This might suggest that early treatment of MDD could impact future neuropsychiatric symptomatology and cognitive performance.

## Introduction

Depression in the elderly has been associated with cognitive difficulties(1). The connections between depression and Mild Cognitive Impairment (MCI) has been reported to be frequently co-morbid, and cognitive disorders occurring in the context of depression may not be remitted(2). The persistence of cognitive impairment after depression is of a significant clinical concern, since it increases the risk of converting from MCI to Alzheimer’s disease (AD)(3). Some studies looked at depressive symptoms as risk factors for accelerated cognitive decline. Wilson et al (2002) reported that elderly participants with depressive symptoms and without AD at baseline, had a 19% increased risk of AD after 7 years and the overall annual decline was increased by 24% in these participants(4). In another prospective longitudinal study, Köhler et al. (2010) showed a robust relationship between depressive symptoms and subsequent cognitive decline(5). Clinically meaningful depressive and persistently high depressive symptoms were associated with faster cognitive decline and increased the risk for development of future “Cognitive Impairment, No Dementia”, especially with memory impairment. More recently, similar data were obtained by Oh et al (2020) from a large Korean cohort showing a threefold increased risk of dementia in non-demented, cognitively normal individuals with subsyndromal depression(6). This risk was also increased in chronic or recurrent subsyndromal depression(6). However, the longitudinal study of depressive symptoms before a diagnosis of dementia by Singh-Manoux et al., carried out over a 28-year follow-up, states that only depressive symptoms in late life but not midlife were associated with increased risk for dementia(7). As such, the potential overlap between depression and age-related cognitive decline is still being questioned.

The potential differences in results could be also due to different methods of quantifying depression. Some studies used specific clinical scales, but this is not in line with the clinical definition of depression, known as “major depressive disorder” (MDD) and which can be evoked from a psychiatric perspective(8). MDD is defined by the DSM-V criteria and requires: (1) the presence of at least one of the two symptoms of (I) depressed mood and (ii) loss of interest or pleasure; (2) as well as at least five of the nine symptoms (first two and: (iii) weight or appetite loss or gain, (iv) insomnia or hypersomnia, (v) psychomotor agitation or retardation, (vi) fatigue, (vii) worthlessness, (viii) diminished ability to think or concentrate and (ix) suicidal ideation)(8). MDD has been linked to various cognitive disorders, particularly memory and dysexecutive disorders, in both adults and the elderly. In the latter population, the severity of MDD has been linked to greater difficulties in initiation, as well as to greater instrumental problems in daily life in individuals without dementia(9). These data were confirmed by Gildengers et al. (2012) who describe dysexecutive impairments and slower processing speed in elderly patients with MDD compared to control participants(10). The presence of cognitive impairments was furthermore not specific to acute phases of MDD but could be found in remittent phases(10). Some of these impairments could then be irreversible precursors of mild cognitive impairment (MCI). Interestingly, MDD has been reported in 11.2-19.6% of MCI patients based on DSM-V criteria(11,12), in 11.5% of mild AD, 10% of moderate AD and 4.5% of severe AD(13).

Although the increased risk of dementia in the presence of neuropsychiatric symptoms (NPS) has already been reported,(14–16) the comorbidities between MDD and other NPS are scarce. Recent factorial models, studied in the review and meta-analysis by Liew (2019), showed that depression was consistently associated with anxiety and often associated with apathy(17). Other studies showed that depression is frequently comorbid with anxiety, and more so in the elderly than in the young(18,19). Depression was also shown to be associated with apathy as well cognitive changes in the form poorer episodic memory and diminished processing speed performance(20). In patients with dementia, depression has been associated with various NPS factors such as hallucinations and anxiety(21). Nevertheless, previous studies concentrated on depression based on clinical scales, and, to our knowledge, the potential associations with the MDD diagnosis have not been studied yet.

Considering the affective (anxiety, apathy) and cognitive (memory and executive functions) associations of MDD in aging and the possible associations with pathological cognitive decline (MCI and AD), as well as the reduction in the prevalence of MDD with increasing severity of decline, this study aims to determine the probability of NPS based on MDD as well as associations of MDD with cognitive performance at different stages of cognitive performance (CH, MCI and AD with MDD compared to those without).

## Methods

### Participants

Data used in the preparation of this article were obtained from the NACC database (specifically NACC UDSv1-2) data. The NACC database is a large compilation of longitudinal data of participants that are CH as well as patients with MCI, AD and other neurodegenerative disorders. Each participant benefited from a neuropsychiatric assessment via the Neuropsychiatric Inventory and a comprehensive cognitive examination. Excluding criteria were: (i) incomplete assessments, (ii) incomplete neuropsychiatric and cognitive assessments, (iii) presence of psychiatric history other than MDD (schizophrenia, bipolar disorder, substance abuse, post-traumatic stress, obsessive-compulsive disorder), (iv) presence of neurological history (stroke, head injury, brain tumor, anoxia, epilepsy, alcohol dependence and Korsakoff, neurodevelopmental disorder), (v) prematurity, (vi) diagnostic criteria in favor of other neurodegenerative or neurological etiology (Parkinson’s disease, frontotemporal degeneration, progressive supranuclear paralysis, corticobasal degeneration, Lewy body dementia, amyotrophic lateral sclerosis, multiple sclerosis, multi-system atrophy, vascular dementia), (vii) presence of substance abuse. After the exclusion, the final sample for analyses consisted of 647 Healthy Controls (CH), 291 patients with MCI and 208 with AD. In the NACC database, patients with AD met the NINCDS/ADRDA criteria for probable or possible AD. Patients with MCI presented a diminished cognitive performance in at least one cognitive domain.

Psychiatric conditions (such as diagnosis and treatment by a physician) are collected at the initial NACC data collection visit. The patients are considered with MDD if the diagnosis of MDD as described by the DSM criteria was attested by a physician in the previous two years. The diagnosis of MDD requires a distinct change of mood, often characterized by sadness and anhedonia accompanied by at least several psychophysiological changes, such as disturbances in sleep, appetite, or sexual desire, cognitive disorders, suicidal thoughts, weak self-esteem, and slowing of speech and action. These changes must last a minimum of 2 weeks and interfere considerably with work and family relations. All participants were further divided in 2 groups based on the presence of MDD in 6 groups: CH-no-MDD, CH+MDD, MCI-no-MDD, MCI+MDD, AD-no-MDD and AD+MDD (Table 1). This study was approved by the Comité d’éthique de la recherche vieillissement-neuroimagerie CER VN 19-20-06. Ethics committee approval and individual patient consents were received by the NACC databases (https://naccdata.org/data-collection/forms-documentation/uds-3).

**Table 1:**
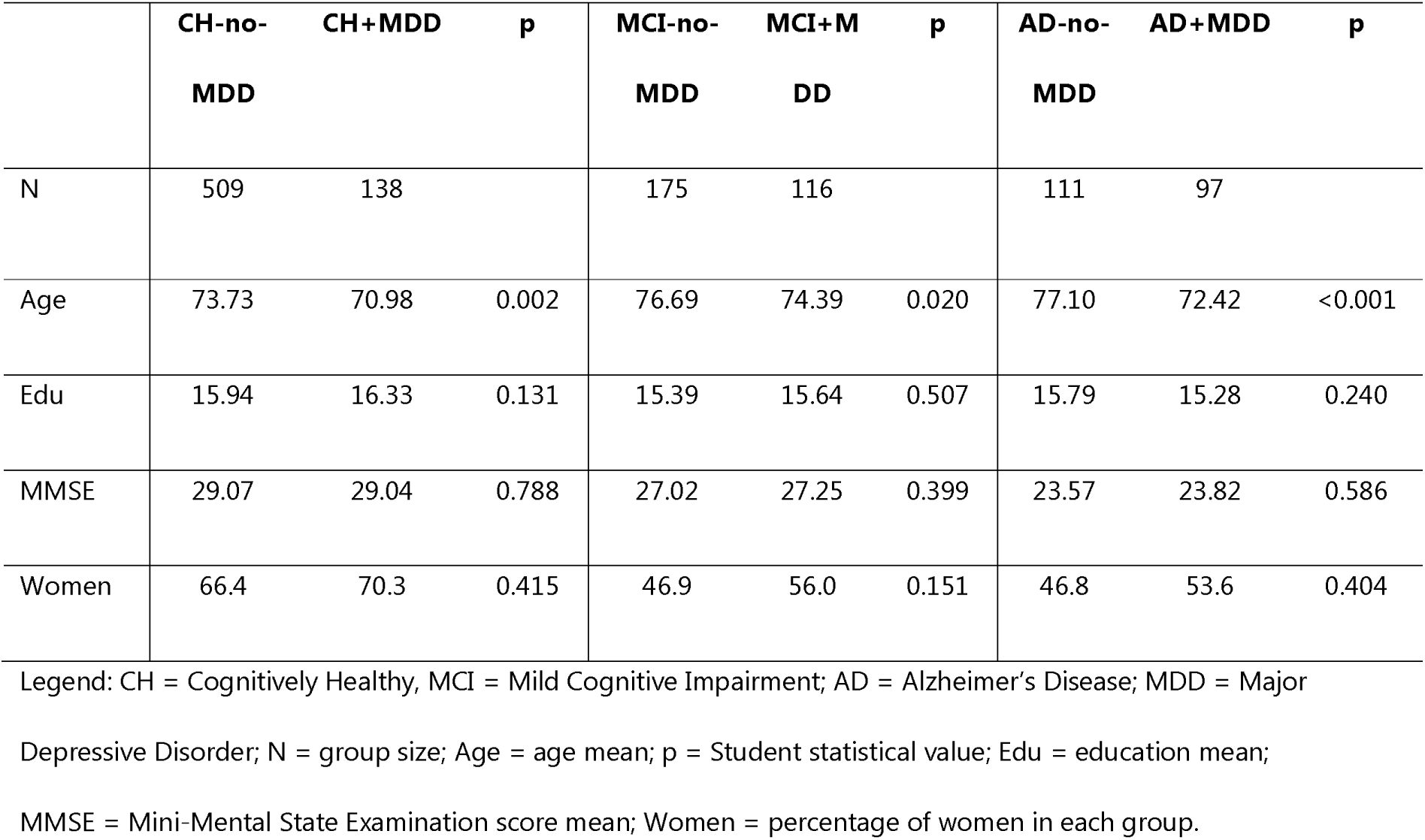
Demographic comparisons between depressive status subgroups for each clinical status.

### Neuropsychiatric and Cognitive Assessments

The NPS were quantified based on the Neuropsychiatric Inventory; version evaluated by participants’ relatives, that was collected during the initial visit. We extracted the values of “presence” / “absence” of 12 NPS: delusions, hallucinations, agitation/aggressiveness, depression, anxiety, euphoria, apathy, disinhibition, irritability, aberrant motor behaviors, sleep disorders and eating disorders.

The cognitive assessment included the following tests present in both datasets: (1) global cognitive efficiency (MMSE), (2) auditivo-verbal memory (logical memory test from Wechsler Memory Scale), (3) focused attention (Trail Making Test A), (4) processing speed (WAIS Coding), (5) mental flexibility (Trail Making Test B), (6) working memory (digit span), (7) semantic verbal fluency (animal and vegetable) and (8) oral naming (Boston Naming Test).

### Statistical analysis

Independent variables were the (1) clinical group (CH or MCI or AD) and (2) MDD presence (+MDD or no-MDD). Demographic data analyses included: (1) a between groups distribution using the student t test for age, years of education, MMSE score, and sex (Table 1) as well as (2) analysis of prevalence of NPS among subgroups as well inter-group differences (Table 2).

**Table 2:**
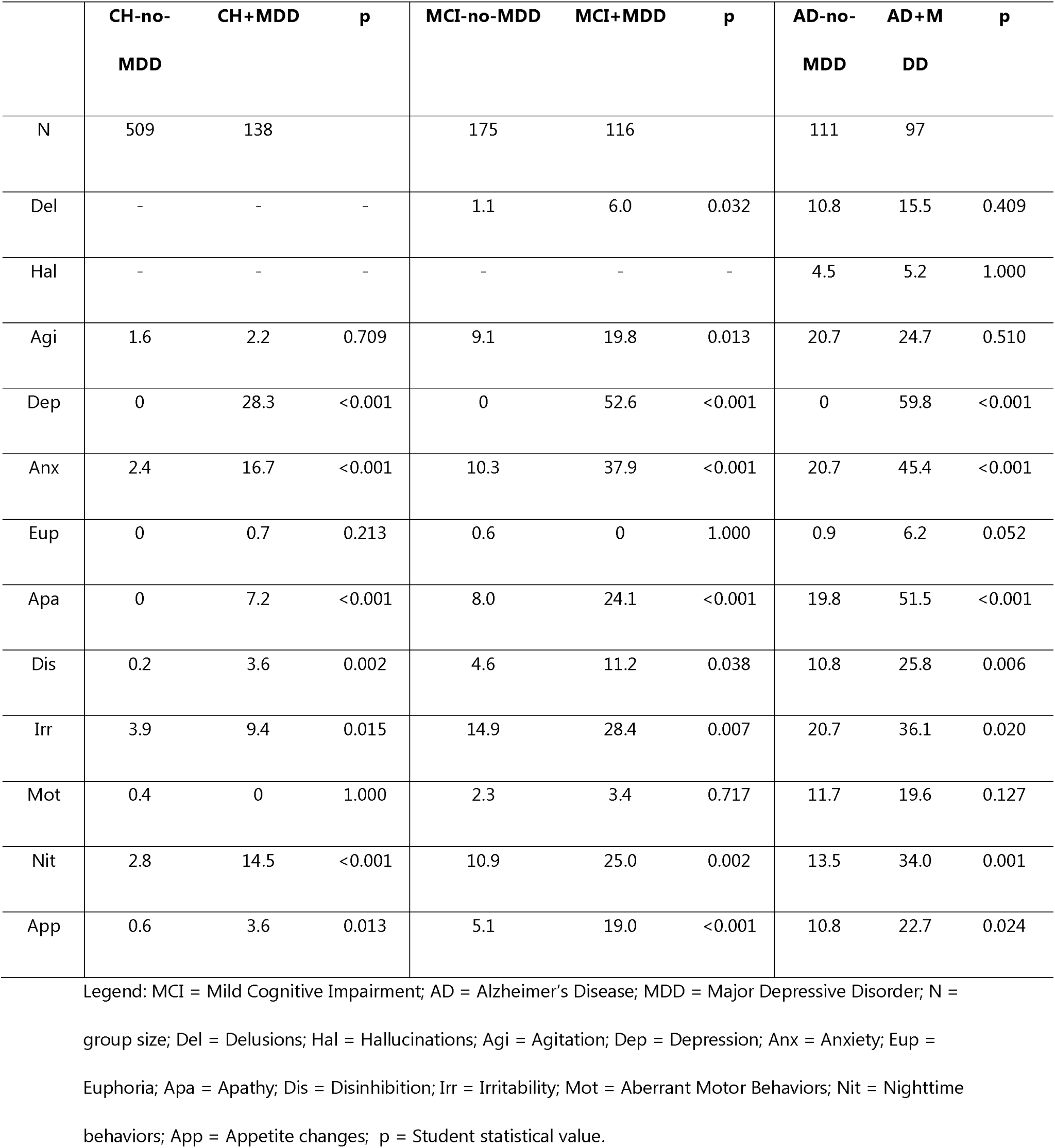
Neuropsychiatric Symptoms prevalence (%) comparisons between depressive status subgroup for each clinical status.

To determine the role of MDD in in the increase the other NPS probability in CH, MCI and AD, we performed binary logistic regression models for each NPS as dependent variables and the presence/absence of MDD as the independent variable.

In a second step, cognitive performance was compared between CH, MCI and AD patients with MDD versus those without MDD using a MANCOVA model with age, sex, and education as covariables (because the +MDD subgroups were younger). Interpretations were made with an alpha threshold of 0.05, and a Bonferroni correction was made for multiple comparisons.

## Results

Demographic, neuropsychiatric and cognitive performance data (Tables 1, 2, 3) showed that the years of education, sex distributions and MMSE score were similar between subgroups within each clinical group. For age, the +MDD subgroups were younger than the no-MDD within each clinical group (Table 1).

**Table 3:** Probability models of NPS based on MDD in CH, MCI and AD.

|  | CH |  |  |  | MCI |  |  |  | AD |  |  |  |
| --- | --- | --- | --- | --- | --- | --- | --- | --- | --- | --- | --- | --- |
| NPS | $\beta$ | p | OR | 95%CI | $\beta$ | p | OR | 95%CI | $\beta$ | p | OR | 95%CI |
| Del | - | - | - | - | 1.715 | 0.035 | 5.555 | 1.133;<br>27.231 | - | - | - | - |
| Hal | - | - | - | - | - | - | - | - | - | - | - | - |
| Agi | - | - | - | - | 0.899 | 0.010 | 2.458 | 1.236;<br>4.887 | - | - | - | - |
| Dep | 20.271 | 0.991 | NS | NS | 21.306 | 0.994 | NS | NS | 21.600 | 0.995 | NS | NS |
| Anx | 2.114 | <0.001 | 8.283 | 4.005;<br>17.134 | 1.673 | <0.001 | 5.330 | 2.881;<br>9.862 | 1.156 | <0.001 | 3.176 | 1.728;<br>5.838 |
| Eup | - | - | - | - | - | - | - | - | 1.981 | 0.069 | 7.252 | 0.857;<br>61.35 |
| Apa | 18.653 | 0.992 | NS | NS | 1.297 | <0.001 | 3.659 | 1.831;<br>7.311 | 1.459 | <0.001 | 4.304 | 2.330;<br>7.948 |
| Dis | 2.950 | 0.007 | 19.098 | 2.212;<br>164.9 | 0.969 | 0.038 | 2.635 | 1.056;<br>6.574 | 1.052 | 0.006 | 2.865 | 1.350;<br>6.078 |
| Irr | 0.933 | 0.012 | 2.543 | 1.231;<br>5.252 | 0.824 | 0.005 | 2.278 | 1.276;<br>4.069 | 0.770 | 0.015 | 2.160 | 1.164;<br>4.009 |
| Mot | - | - | - | - | - | - | - | - | - | - | - | - |
| Nit | 1.791 | <0.001 | 5.993 | 2.941;<br>12.21 | 1.007 | 0.002 | 2.737 | 1.450;<br>5.165 | 1.194 | 0.001 | 3.300 | 1.659;<br>6.562 |
| App | 1.847 | 0.012 | 6.341 | 1.496;<br>26.87 | 1.463 | <0.001 | 4.317 | 1.909;<br>9.760 | 0.884 | 0.024 | 2.420 | 1.126;<br>5.199 |

Legend: CH = Cognitively Healthy; MCI = Mild Cognitive Impairment; AD = Alzheimer's Disease; NPS = Neuropsychiatric Symptoms; $\beta$ = coefficient of regression models. A positive coefficient indicates that presence of MDD increase the probability of NPS; p = p-value; OR = Odd Ratio; Del = Delusions; Hal = Hallucinations; Agi = Agitation; Dep = Depression; Anx = Anxiety; Eup = Euphoria; Apa = Apathy; Dis = Disinhibition; Irr = Irritability; Mot = Aberrant Motor Behaviors; Nit = Nighttime behaviors; App = Appetite changes; empty values = prevalence of NPS is insufficient for analysis or the independent factor does not significantly change the model based on the constant alone; NS = not significant.

Prevalence analysis showed that within the two groups MCI and AD, those with MDD exhibited more frequently anxiety, apathy, disinhibition, irritability, disturbed nighttime behavior, and appetite changes than those without MDD. Furthermore, MCI+MDD participants had more delusions and agitation than MCI-no-MDD. Within the group CH, those with MDD exhibited more frequently anxiety, apathy, disinhibition, irritability, nighttime behaviors and appetite changes (Table 2).

### Neuropsychiatric associations of MDD

In the CH group, the presence of MDD was positively and significantly involved in the probability equations for anxiety, disinhibition, irritability, nighttime behaviors, and appetite changes. However, MDD alone does not appear to improve the model sensitivity (Specificity = 100%, sensitivity = 0%). In the MCI group, the MDD was positively and significantly involved in the probability equations for delusions, agitation, anxiety, apathy, disinhibition, irritability, nighttime behaviors, and appetite changes. Thus, the presence of MDD increased the likelihood of experiencing these NPS. However, MDD alone does not appear to improve the model sensitivity in MCI (Specificity = 100%, sensitivity = 0%).

In the AD group, the MDD was positively and significantly involved in the probability equations for anxiety, euphoria, apathy, disinhibition, irritability, nocturnal behaviors, and appetite changes. In AD, MDD improve the model sensitivity for apathy only (specificity = 65.4%, sensitivity = 69.4%).

### Cognitive association of MDD

After controlling for covariates, the MANCOVA model showed no cognitive associations with MDD in CH participants: mean performance was similar between CH+MDD and CH-no-MDD.

Among participants with MCI, MCI+MDD performed worse on the Trail Making Test A and B (TMTA and TMTB), with longer completion times than MCI-no-MDD, reflecting poorer performance in focused attention and mental flexibility respectively.

Finally, among participants with AD, AD+MDD performed better on the digit span backward task (DSB), reflecting a better auditory-verbal working memory compared to AD-no-MDD patients (Table 4).

**Table 4:** Comparisons of cognitive performance means between depressive status subgroups for each clinical status (Controlling for age, sex, and education as covariables).

| Variable | CH |  |  | MCI |  |  | AD |  |  |
| --- | --- | --- | --- | --- | --- | --- | --- | --- | --- |
|  | Differenc<br>e<br>MDD –<br>no-MDD | F | p | Differenc<br>e<br>MDD –<br>no-MDD | F | p | Differenc<br>e<br>MDD –<br>no-MDD | F | p |
| LM | -0.025 | 0.005 | 0.945 | -0.056 | 0.011 | 0.916 | -0.053 | 0.010 | 0.919 |
| LMD | -0.066 | 0.027 | 0.868 | -0.765 | 1.898 | 0.169 | 0.042 | 0.007 | 0.934 |
| DSF | 0.037 | 0.038 | 0.846 | 0.275 | 1.079 | 0.300 | 0.509 | 2.926 | 0.089 |
| DSB | 0.129 | 0.381 | 0.537 | 0.427 | 3.076 | 0.081 | 0.756 | 6.973 | 0.009 |
| Ani | -0.165 | 0.109 | 0.742 | 0.095 | 0.026 | 0.873 | -0.148 | 0.041 | 0.839 |
| Veg | -0.450 | 1.480 | 0.224 | 0.177 | 0.162 | 0.688 | 0.828 | 2.916 | 0.089 |
| TMTA | 0.147 | 0.019 | 0.890 | 5.824 | 7.373 | 0.007 | 5.375 | 1.345 | 0.247 |
| TMTB | 3.621 | 1.036 | 0.309 | 19.285 | 5.252 | 0.023 | 13.787 | 1.268 | 0.261 |
| Code | -0.162 | 0.027 | 0.869 | -2.276 | 3.744 | 0.054 | -2.453 | 1.700 | 0.194 |
| BNT | -0.283 | 1.033 | 0.310 | 0.002 | <0.001 | 0.996 | 0.967 | 1.234 | 0.268 |

Legend: MCI = Mild Cognitive Impairment; AD = Alzheimer's Disease; MDD = Major Depressive Disorder; F = ANCOVA statistical value; LM = Logical Memory immediate recall; LMD = Logical Memory Delayed recall; DSF/DSB = Digit Span Forward/Backward; Ani = Animal verbal fluency; Veg = Vegetables verbal fluency; TMTA = Trail Making Test version A; TMTB = Trail Making Test version B; Code = WAIS code subtest; BNT = Boston Naming Test; p = Student statistical value.

## Discussion

When considering 2 levels of cognitive decline: MCI and AD, the regression models and analysis of covariance performed in our study, showed a role of MDD in increase of the NPS probability and a significant impact of MDD on executive performances. These results remained significant even when the MANCOVA model included that confounding factors of age, sex, and years of education. The presence of MDD is associated with a higher prevalence of delusions and agitation in the MCI group and more prevalence of anxiety, apathy, disinhibition, irritability, disturbed nighttime behaviors and appetite disturbance in the MCI and AD groups. Moreover, logistic regression models confirm these associations, MDD being a significant involvement in determining the presence of these NPS, implying that a patient with MDD had a higher probability of having an additional NPS. In CH participants, MDD was only neuropsychiatric impact, featured by higher prevalence of anxiety, disinhibition, irritability, nighttime behaviors and appetite changes.

These relationships might suggest a likely mood lability in MCI and AD patients, as mood can shift from apathy to aggressivity or from depression to euphoria in these patients. Previous studies outlined this link by describing that emotional lability can be frequent in AD patients, as well as the emergence of late onset “bipolar disorder” type syndromes with manic episodes(22). Moreover, the emergence of these disorders has been described as a risk factor in the development of dementia(23). Interestingly, previous studies also demonstrated an association between depression and disinhibition in MCI and AD, though it was less frequently evidenced(24).

The significance of agitation, depicted by our model, is in line with previous literature. Agitation was commonly described in MCI and AD and appeared to be associated with most other neuropsychiatric and behavioral disorders, including depression, even in MCI(24–26). Agitation has been shown to be an expression of anxiety in AD patients(27). In contrast, depression and apathy appeared to be more closely associated with the incidence of agitation and the agitation composite score (which includes anxiety, agitation, irritability, disinhibition, and aberrant motor behavior) as reported by Liu et. al. 2020(28).

Our study showed no relation between MDD and hallucinations, whereas several previous works reported them in AD, specifically with respect to visual, auditory, and olfactory hallucinations(29–30). Thus, it could be that the occurrence of hallucinations is not directly due to depression but mediated by it. These may also be accentuated by social isolation, loneliness and sensory deprivation(31–32). On the other hand, behaviors of euphoria or excessive joviality have also been described in the context of late bipolarity(22).

Overall, our results show that patients with MDD had more frontal-type symptoms (such as disinhibition, agitation or irritability) and no aberrant motor behaviors. Even if previous literature described motor behaviors in relation to frontal-type symptoms, our study shows a potential distinct impact of MDD on this NPS(24). However, these behaviors are poorly studied and do not seem to have direct links with depression(33). According to Martin & Velayudhan’s (2020) literature review on NPS in MCI, some symptoms that are less frequent, or even as frequent as in the general population, would not present a higher risk factor for developing a dementia state or accelerated cognitive decline compared to people without these NPS: these are disinhibition, euphoria, appetite/eating disorders, and aberrant motor behavior(34). According to this, a recent study looked at clusters of NPS in MCI and their comparative risks of dementia and found that frontal/hyperactivity-like symptoms, such as euphoria, disinhibition, and aberrant motor behaviors would not present an increased risk for patients to develop dementia, and that these symptoms would be a consequence of other symptoms such as psychotic or affective symptoms(17).

Our results also showed that the MCI+MDD group was characterized by poorer performance on selective attention and mental flexibility in comparison to the MCI-no-MDD group while the AD+MDD was characterized by better auditivo-verbal working memory performance. This would imply that MDD potentiates reductions in cognitive performance in MCI but not in AD. This effect is too challenging to explain at this stage. Some studies have shown a protective role of certain antidepressants against cognitive decline (dementia or MCI)(35). Thus, treatment of MDD may provide compensatory protection from cognitive decline. However, these results are not systematically confirmed and only include certain treatments, such as tricyclic antidepressants or lithium, with other antidepressants tending to increase the risk of dementia, added to the accentuated risk due to depression(36).

Executive dysfunction has been described in AD and associated with some NPS such as depression(29). It has been suggested that depression is a risk factor for dysexecutive disorders in AD. But these results are not in line with our results and other studies showing an impact of depression on executive function in MCI but not in AD(37). It seems that in AD patients, severe depressive symptoms may play an important role in cognitive impairment, while less severe depressive symptoms may have limited effects(37). Other studies have linked depression with dysexecutive disorders as well as the severity of depression as a mediator of the functional impact of executive disorders(38). Here, a limitation could be that the reduction of executive functions in MCI was not totally explained by the presence of depression due to subtypes of MCI not considered in this study. Indeed, some subtypes can show more severe dysexecutive disorders without high prevalence of depression while other subtypes show higher prevalence of depression with less cognitive impairment(39). Further investigations are required to consider other impact factors such as the duration of depressive symptoms or their frequency.

In connection with the debate about depression as a risk factor or prodromal sign of AD and dementia, Köhler et al (2010) recalled that there is evidence that depression may accompany pre-clinical AD, or precede the onset of AD by several years, in the sense that depressive episodes in the past life increase the risk of subsequent AD(5). Thus, depressive symptoms in the broad sense may have this dual role of risk factor and consequence of AD.

Our study should be viewed in light on some limitations: (1) In the analyses, the presence or absence of treatments for MDD was not considered; (2) some patients also had earlier MDD that was treated, these patients were not included in the study due to a much smaller sample size.

In conclusion, our study shows associations between MDD and a higher prevalence of NPS, while the relationship with cognitive performance were weak. It is important that future studies pay more attention to the characteristics of the processes that are broadly referred as “depression,” which may both involve a clinical picture of a mental disorder or more isolated symptoms occurring in the course of aging.

## Data Availability

All data are available on the NACC website upon demand (https://naccdata.org/requesting-data/submit-data-request).

## AUTHORS’ CONTRIBUTIONS

Research project: AH, LR; Data extraction and processing: LR; Statistical analysis: LR; Manuscript: LR, AH

## Use of NACC data

*The NACC database is funded by NIA/NIH Grant U01 AG016976. NACC data are contributed by the NIA-funded ADCs: P30 AG019610 (PI Eric Reiman, MD), P30 AG013846 (PI Neil Kowall, MD), P50 AG008702 (PI Scott Small, MD), P50 AG025688 (PI Allan Levey, MD, PhD), P50 AG047266 (PI Todd Golde, MD, PhD), P30 AG010133 (PI Andrew Saykin, PsyD), P50 AG005146 (PI Marilyn Albert, PhD), P50 AG005134 (PI Bradley Hyman, MD, PhD), P50 AG016574 (PI Ronald Petersen, MD, PhD), P50 AG005138 (PI Mary Sano, PhD), P30 AG008051 (PI Thomas Wisniewski, MD), P30 AG013854 (PI Robert Vassar, PhD), P30 AG008017 (PI Jeffrey Kaye, MD), P30 AG010161 (PI David Bennett, MD), P50 AG047366 (PI Victor Henderson, MD, MS), P30 AG010129 (PI Charles DeCarli, MD), P50 AG016573 (PI Frank LaFerla, PhD), P50 AG005131 (PI James Brewer, MD, PhD), P50 AG023501 (PI Bruce Miller, MD), P30 AG035982 (PI Russell Swerdlow, MD), P30 AG028383 (PI Linda Van Eldik, PhD), P30 AG053760 (PI Henry Paulson, MD, PhD), P30 AG010124 (PI John Trojanowski, MD, PhD), P50 AG005133 (PI Oscar Lopez, MD), P50 AG005142 (PI Helena Chui, MD), P30 AG012300 (PI Roger Rosenberg, MD), P30 AG049638 (PI Suzanne Craft, PhD), P50 AG005136 (PI Thomas Grabowski, MD), P50 AG033514 (PI Sanjay Asthana, MD, FRCP), P50 AG005681 (PI John Morris, MD), P50 AG047270 (PI Stephen Strittmatter, MD, PhD). All data are available on the NACC website upon demand (https://naccdata.org/requesting-data/submit-data-request)*.

